# Surveying the Literature on Implementation Determinants and Strategies for HIV Structural Interventions: A Systematic Review Protocol

**DOI:** 10.1101/2025.01.02.25319901

**Authors:** alithia zamantakis, Shruti Chandra, Valeria A. Donoso, R. Mariajose Paton, Alec Powers, Brian Mustanski, Nanette Benbow

**Author notes:** Corresponding Author: Nanette Benbow.

## Abstract

**Background:** Despite improvements in HIV prevention, treatment, and surveillance, vast disparities remain in access, uptake, and adherence of evidence-based interventions. These disparities are most pronounced among racially, sexually, and gender minoritized populations, as well as among those living in poverty and/or who use injectable drugs. Structural interventions, or interventions that target social and structural determinants of health like housing, transportation, or income, are needed to increase access to, use of, and adherence to HIV EBIs to advance the aims of the national Ending the HIV Epidemic initiative. However, it is unclear to what extent barriers and facilitators of structural interventions have been identified in the U.S. and what implementation strategies and adjunctive interventions have been developed to enhance their delivery.

**Methods:** To identify what implementation determinants, implementation strategies, and adjunctive interventions have been identified for HIV structural interventions, we carried out a broad database search between May and July 2024, identifying a total of 8,098 articles. We will use a multi-step process to identify articles to include in the systematic review. We will use natural language processing to identify articles for exclusion, followed by manual text review and extraction using COVIDENCE software. Literature on determinants will be coded according to the Consolidated Framework for Implementation Research. Implementation strategies and adjunctive interventions will be coded according to the Expert Recommendations for Implementing Change, the Theoretical Domains Framework, and COM-B. We will descriptively analyze determinants, implementation strategies, and adjunctive interventions, use natural language processing for thematic analysis of determinants, implementation strategies, and adjunctive interventions, and provide narrative description of implementation strategies and adjunctive interventions.

**Discussion:** This systematic review will identify key barriers and facilitators for HIV structural intervention implementation strategies, including multi-level approaches to address disparities among marginalized populations. Findings will provide insights for advancing equitable, scalable interventions to support the goals of the Ending the HIV Epidemic initiative.

**Systematic review registration:** CRD42024554315

## Background

The HIV epidemic disproportionately impacts minoritized populations, underscoring the need to examine the social and structural determinants of health (SSDOH) that contribute to these inequities (1,2). SSDOH include the “upstream” or structural factors like public policy, economic context, social and cultural values, and political ideology that produce or give shape to social determinants of health (3). SSDOH, thus, also include the more “mid-stream” or social factors, often referred to as the conditions where individuals work, live, grow, and play, that shape a populations’ health before individuals step foot in a clinic (3). The federal government has identified “key populations” that are disproportionately affected by the HIV epidemic due, in part, to SSDOH: cisgender gay, bisexual, and other men who have sex with men of all races and ethnicities (MSM); Black women people who inject drugs (PWID); youth aged 13 to 24 years; and transgender women (4). Despite improvements in overall HIV incidence due to the development and implementation of numerous evidence-based interventions (EBIs), trends in the United States highlight the disparities between these groups and the general population (5). For example, between 2018 and 2022, new HIV diagnoses in Black, Indigenous, Hispanic/Latine, and Native Hawaiian or Pacific Islander populations increased, but decreased or stabilized in Asian and White populations, respectively (5). SSDOH such as racism, access to care, economic status, and social stigma can all contribute to discrepancies in HIV diagnosis, treatment, and prevention rates (6).

Pre-exposure prophylaxis (PrEP), an oral pill or bimonthly injection that significantly reduces the risk of acquiring HIV (7–9), is a critical tool in HIV prevention, but disparities in access and uptake present significant challenges. For example, PrEP uptake has been slow among Black and Latine individuals, particularly Black females and Latino males (10,11). Barriers such as stigma, medical mistrust, and structural racism contribute to the inequitable distribution of PrEP, limiting its full potential in reducing HIV incidence (12).

Treatment as Prevention, another critical innovation, involves the suppression of individuals’ HIV viral load through consistent antiretroviral therapy (ART) adherence and has emerged as a highly effective strategy in preventing the transmission of HIV through sex, syringe sharing, and vertical (mother-to-child) transmission (13–18). However, barriers to access to care and to ART adherence remain prevalent among key populations, including housing instability, lack of access to transportation, substance use, which each, without further patient support, contribute to patients missing a greater number of clinic appointments (19).

Achieving the goals of the Ending the HIV Epidemic in the US (EHE) Initiative to reduce HIV incidence and improve health equity necessitates the implementation of structural interventions (20). Structural interventions attempt to make changes to the performance and creation of public health initiatives (21). These interventions target the structural factors of a public health initiative that can influence the target population’s behavior. The structural factors (or determinants) influencing health behaviors can include social inequalities, economic factors, cultural influences, policies, and physical structures (21). Structural interventions have been used as a preventative strategy to target the structural variables that impact behavior and risk of obtaining HIV. For example, the provision of housing as an intervention has been found to reduce engagement in sexual and substance use behaviors that may increase the likelihood of acquiring HIV, as well as to increase access and adherence to ART (22).

While a growing body of literature has attended to developing and piloting structural interventions, there has been less attention to their implementation within the U.S. Implementation science has identified a persistent gap between the development, piloting, and trialing of an EBI and its eventual implementation (referred to as the implementation gap (23)). Thus, it is necessary for researchers to identify the implementation determinants, or barriers and facilitators, of structural interventions and the implementation strategies or processes, methods, policies, and programs to increase the reach, effectiveness, adoption, implementation, and sustainability of the interventions (24–27). Structural interventions will also likely require the identification and development of adjunctive interventions, or methods to increase patient uptake, access, and adherence to the intervention (28). Implementation science’s focus on swift translation of research to practice is necessary to achieve the goals of the EHE initiative (29).

Thus, we aim to carry out a systematic review to identify:

1. What implementation determinants, implementation strategies, and adjunctive interventions have been identified as relevant to the adoption, acceptability, feasibility, appropriateness, reach, cost, sustainability, and equitable delivery of structural interventions for HIV EBIs?
2. What gaps are present in the literature regarding the implementation of structural interventions for HIV EBIs?
3. How are or how may structural interventions be differentiated from implementation strategies and adjunctive interventions?

## Methods

### Search Strategy

Between May and July 2024, a broad database search strategy (see **Supplemental File 1**) was carried out to identify literature focused on implementation determinants, implementation strategies, and adjunctive interventions for structural interventions for HIV EBIs along the HIV prevention and care continua (30,31). The protocol for this search is registered with the International Prospective Register of Systematic Reviews (PROSPERO ID: CRD42024554315). A research librarian provided consultation on the search strategy and recommended additional databases to search. The first author (az) searched Ovid MEDLINE [2000-2024], Embase (Elsevier) [2000-2024], PsycINFO (EbscoHost) [2000-2024], Web of Science [2000-2024], Sociological Abstracts [2000-2024], Google [All years], and ProQuest Congressional CRS Reports and Miscellaneous Publications [2000-2024]. The second author (VAD) also identified the primary sources from the CDC’s compendium of evidence based structural interventions for HIV to add to the initial search (32). From this, 8,098 articles were identified before duplication. Using these articles, we will conduct a systematic review in two phases: 1) identifying implementation determinants and 2) identifying implementation strategies and adjunctive interventions. Both phases of the systematic review will be carried out in compliance with the Preferred Reporting Items for Systematic Reviews and Meta-Analyses (PRISMA; see **Supplemental File 2**).

### Screening and eligibility criteria

We used a multi-step process to identify articles to include in the systematic review. Inclusion criteria include: 1) a focus on HIV/AIDS; 2) 50% or more of the study sample is based in the U.S.; 3) the study is an implementation or hybrid implementation-effectiveness study (33); 4) the study contains original research with an original data sample; 5) behavioral or social science (i.e., not basic science); 6) the study reports, explores, or tests at least one implementation outcome (i.e., acceptability, feasibility, adoption, fidelity, implementation costs, reach, sustainability (26,27)); and 7) that the intervention either (a) affects risks and/or behavioral choices by changing something external to the individual and not under their control and/or (b) targets social determinants of health.

Preliminary inclusion was determined using natural language processing (NLP) to analyze titles and abstracts (N = 8,092) and classify articles for eligibility in our systematic review. NLP was performed by author MH using R version 4.4.1 (34) and packages quanteda for quantitative analysis of textual data (35), RWeka for machine learning algorithms for data mining (36), and tidyverse (37), tidytext (38), broom (39), readr (40), stringr (41), and magrittr (42) for data cleaning and manipulation. Five domains, with separate R scripts, were defined: 1) Domestic Setting (i.e., not international or foreign); 2) Study Design; 3) Behavioral Science; 4) HIV/AIDS Relevance; and 5) Structural Intervention. Using the quanteda package in R, each article was assigned a label for inclusion, exclusion, further inspection required, or no label based on the quantified presence of predetermined terms. 405 articles were assigned an ‘include’ label across all five domain classifiers, and 4,285 articles contained at least one ‘further inspection’ label and were not assigned an ‘exclude’ label across any classifier. Lastly, results were output into a table indicating the classification of each article.

After computerized exclusion, articles identified for inclusion in the NLP were uploaded to COVIDENCE for manual review (43). COVIDENCE will automatically identify duplicate articles for exclusion. A team of trained investigators, including but not limited to a faculty investigator with experience leading systematic reviews (az), a postdoctoral fellow (MP), and two research staff with Bachelor-level training (VAD, SC), will conduct the manual review. The research staff and postdoctoral fellow were trained by az to review articles until 90% reliability was reached between their identification of articles to include and az’s identification of articles to include. Manual review will begin with title and abstract review. The team of investigators will screen each article’s title and abstract to identify whether to include or exclude each article based on the same criteria as the NLP will use. Following, title and abstract review, we will conduct full-text review of each article, once again screening each article to identify whether to include or exclude each article based on the same criteria as the prior two steps. In both steps of manual screening, each article will be reviewed by two investigators. In the case of disagreement, the article will be re-reviewed by senior members of the team (az and NB) for consensus.

### Extraction

Following the multi-step screening process, we will manage and extract data from articles using COVIDENCE software. Extraction will include identification of high-level information detailed in Table 1. This will include CDC-identified priority populations, which are based on disparities in HIV prevalence and incidence in the U.S. Study type will be categorized as within-site designs (e.g., post-design examination of healthcare processes), between-site designs (e.g., comparing a new implementation strategy to usual practice or comparing different implementation strategies), within- and between-site designs (e.g., roll-out designs), non-experimental designs (e.g., qualitative designs, surveys that aren’t part of a larger trial), configurational comparative methods (e.g., qualitative comparative analysis, coincidence analysis), simulation study, or other type of design (44). Implementation outcomes, or “the effects of deliberate and purposive actions to implement new treatments, practices, and services,” will be categorized according to the oft-cited Proctor et al. (2011) framework for implementation outcomes, which includes acceptability, adoption, appropriateness, costs, feasibility, fidelity, penetration, and sustainability (26). In addition to other extraction variables, we will categorize structural intervention types using a taxonomy developed by Sipe et al. (2017). These include interventions aimed at increasing access to an HIV EBI, policy/procedure interventions, the use of mass media to disseminate an intervention, interventions changing the physical structure where an HIV EBI is delivered, capacity-building interventions, community mobilization as an intervention, and interventions targeting SDOH (45). Extraction will be conducted by two members of the investigative team (VAD, MP, SC) for each article. Consensus for any discrepancies will be reviewed for final decision making by senior members of the team (az, NB).

**Table 1.** Information to be extracted from articles included in the review.

|  |  |
| --- | --- |
| Item | Definition/Explanation |
| Article Title | -- |
| Publication Year | -- |
| First Author's Name | -- |
| Journal Name | -- |
| Geographic Setting | Northeast, West, Midwest, South, U.S. colony, nationwide, simulation, unspecified, or other |
| Inner Setting<sup>1</sup> Verbatim | Where the intervention is being delivered (e.g., HIV clinic, mobile testing unit) |
| Inner Setting Category | Specialized HIV Clinic, Substance Use Treatment Facility, Non-Specialized Private Clinic or Hospital, Community Health Centers, Pharmacy, Dental Clinic, Hospital System, Community Organization, Church, Prison/Jail, Emergency Department, Labor Unit/Department, Community Level, Legislative Bodies |
| Unit(s) of Analysis | Patients, providers, policy, non-primary data |
| Patient Population | If applicable; Who the paper identifies as their patient population |
| CDC Priority Population | Cisgender, gay and bisexual Men, African Americans, persons who use drugs, Latines, transgender individuals, cisgender women, adolescents (13-25) |
| Provider Role Verbatim | If applicable; Which role the paper identifies as delivering the intervention (e.g., HIV tester) |
| Provider Role Category | If applicable; HIV Tester, Physician, Pharmacist, Nurse, Laboratory Personnel, Community Health Worker, Program Leadership/Administration, Administrative Assistant, Policymaker, Peer Navigator, Case Manager, Business Owner, Other |
| Higher Level Unit of Analysis | If applicable (e.g., Policymakers, Public Health Department, Nonprofit Organization, Private Entities, Community) |
| HIV Evidence-Based Intervention | PrEP, HIV testing/diagnosis, Linkage to HIV Care, HIV Treatment, nPEP<sup>2</sup>, or other intervention |
| Data Type | Qualitative, Quantitative, Mixed/Multi-Method, Simulation, N/A or Not Reported, and Other |
| Data Collection Method | Electronic Health/Medical Record, Survey, Surveillance, Participant Observation, Interview, Focus Group, Administrative Data, Policy Analysis, Document Review, Simulation, or Other |
| Study Design | Between-Site; Within- and Between-Site; Non-Experimental; Configurational Comparative Methods; Simulation Study; Other |
| Implementation Variables Studied | Determinants Only, Strategies Only, Both Strategies and Determinants |
| Implementation Outcome | Appropriateness, Adoption, Acceptability, Feasibility, Cost, Penetration, Sustainability, Other |
| Implementation Outcome Definition | How do the authors operationalize the implementation outcome |
| Structural Intervention Type | Access, Policy/Procedure, Mass Media, Physical Structure, Capacity-Building, Community Mobilization, Social Determinants of Health Intervention, Other |
| Structural Intervention Outcome | Access, Stigma Reduction, Housing, Food Stability, Poverty Alleviation, Use of Sterilized Syringes, Equity, Other |

<sup>1</sup> Inner setting is a domain of CFIR, which refers to the setting in which an intervention is delivered.
<sup>2</sup> nPEP refers to nonoccupational post-exposure prophylaxis, an oral pill taken within 72 hours of exposure to HIV, which, if taken as prescribed, can prevent HIV infection.

### Determinants Coding

Following extraction, articles identified as assessing implementation determinants will be uploaded to MaxQDA, a qualitative data analysis software, for coding (46). Determinants coding will focus on barriers and facilitators to implementing the structural intervention, rather than barriers and facilitators to implementing an HIV EBI. Determinants will be coded according to valence (i.e., barrier, facilitator, both, neither) and structural intervention type. We will also code the determinant level; that is, whether the determinant is an innovation determinant or an implementation determinant. In comparison to implementation determinants, innovation determinants are barriers or facilitators to patients or other recipients of an EBI access, using, adhering to, or otherwise receiving the intervention (47). Finally, we will code determinants using the domains and constructs of the Consolidated Framework for Implementation Research (CFIR) 2.0 as codes (24,25). CFIR is the most widely-cited compendium (48,49) of implementation determinants and includes five domains: 1) innovation characteristics (i.e., factors about the EBI itself that make it more or less difficult to be delivered); 2) outer setting (i.e., contextual factors in the larger society or community); 3) inner setting (i.e., contextual factors within the setting in which an EBI is delivered that make it more or less difficult to do so); 4) individuals (i.e., whether individuals delivering an EBI have the capability, opportunity, and motivation to do so); and 5) process (i.e., factors about the actual process of implementing an EBI that hinder or facilitate delivery, such as engaging patients or adapting the EBI). The investigative team (VAD, SC, MP) will be trained by a senior team member (az) in use of MaxQDA and coding of determinants. We will continue training and practice coding until an interrater reliability of at least κ = .70 is reached.

### Strategy and adjunctive intervention coding

Following extraction, articles identified as assessing strategies (including implementation strategies and adjunctive interventions) will be coded in Excel. Coding will be performed of implementation strategies and adjunctive interventions for structural interventions, rather than implementation strategies and adjunctive interventions for HIV EBIs. The investigative team (VAD, SC, MP) will be trained by a senior team member (az) in coding of strategies. Each article will be coded by two investigators who will meet to review their individual coding and come to consensus on discrepancies. In the case that they cannot come to consensus, a senior team member (NB) will be brought in to finalize consensus. Strategy coding will begin by identifying whether the article is examining an implementation strategy or an adjunctive intervention.

Implementation strategy coding will include 1) identification of the discrete strategies, their names, their temporal order, and their definition/description; 2) differentiation of the structural intervention and the implementation strategies (if possible); 3) coding of each discrete implementation strategy to Expert Recommendations for Implementing Change (ERIC) framework (50); 4) identification of the specific implementation outcomes each discrete strategy is targeting; 5) identification of the effectiveness of the strategy based on quantitative evidence of effectiveness in comparison to a control group, baseline, or other comparator; 6) whether the investigators assess health disparities and/or health equity, with health disparities defined as identifying a disparity and health equity defined as targeting disparities, including a priority population, and/or having a community-based study design; 7) identification of the stage of implementation research (i.e., are they developing, piloting, testing/trialing, and/or comparing strategies); 7) study design (e.g., observational, within-site); 8) sample size; 9) whether the investigators use a hybrid approach in the study design (i.e., hybrid implementation-effectiveness approach (44)); 8) assessment of how well specified each discrete strategy is according to recommendations by Proctor et al. on a 3-point scale, with each coder rating the level of specification (0 = no details provided; 1 = very little detail; 2 = there is some detail about components of the strategy but key details are left out; 3 = investigators clearly indicate the components of the implementation strategy such that it could be replicated (51)); and 9) whether the authors identified the specific determinants the strategy aims to target.

Adjunctive intervention coding will include 1) identification of the discrete components of the adjunctive intervention (e.g., training, on-going supervision, audit and feedback); 2) description of the adjunctive intervention components; 3) coding the adjunctive intervention components to the Theoretical Domains Framework (TDF) and COM-B (52,53); 4) identification of the specific outcomes each discrete component is targeting (i.e., intervention awareness, acceptability, feasibility, uptake, adherence, costs, and maintenance, implementation strategy acceptability, appropriateness, feasibility, and fidelity); 5) whether the investigators assess health disparities and/or health equity, with health disparities defined as identifying a disparity and health equity defined as targeting disparities, including a priority population, and/or having a community-based study design; 6) identification of the effectiveness of the adjunctive intervention based on quantitative evidence of effectiveness in comparison to a control group, baseline, or other comparator; 7) identification of the stage of implementation research (i.e., are they developing, piloting, testing/trialing, and/or comparing adjunctive interventions); 8) study design (e.g., observational, within-site); 9) sample size; 10) whether the investigators use a hybrid approach in the study design (i.e., hybrid implementation-effectiveness approach); 11) assessment of how well specified each discrete component of the adjunctive intervention is; 12) identification of who the intervention is delivered to; 13) whether the investigators identify the recipient/patient-level determinants the adjunctive intervention aims to target.

### Analysis

#### Determinants

We will export data to Excel to tabulate the number of discrete determinants by CFIR constructs. We will further stratify determinants by delivery setting, CDC priority populations, study method, type of structural intervention (45), the particular HIV EBI the structural intervention is targeting, and implementation outcomes associated with the determinants. Due to the large amount of data potentially analyzed, we will use NLP to thematically analyze determinants to provide narrative description of the types of determinants identified across papers (54).

#### Strategies and adjunctive interventions

For implementation strategies and adjunctive interventions, we will tabulate the number of discrete implementation strategies according to ERIC strategies and the number of adjunctive intervention components according to TDF and COM-B constructs. We will further stratify implementation strategies and adjunctive interventions by study design, stage of research, type of structural intervention (45), the particular HIV EBI the structural intervention is targeting, delivery setting, the patient or provider population the implementation strategy or adjunctive intervention is targeting, and implementation and recipient/patient-level outcomes. We will describe implementation strategy and adjunctive intervention specification by mean specification rating. We will also tabulate the number of studies addressing health disparities and/or health equity. We will tabulate the number of structural interventions that, themselves, are implementation strategies and/or adjunctive interventions. Due to the large amount of data potentially analyzed, we will use NLP to thematically analyze strategies and adjunctive interventions to provide narrative description of the types of strategies and adjunctive interventions identified across papers (54).

### Dissemination

We will disseminate the results of this systematic review via two primary papers: one focused on determinants of structural interventions for HIV EBIs and one focused on implementation strategies and adjunctive interventions for HIV EBIs. We will produce additional qualitative manuscripts examining determinants, implementation strategies, and adjunctive interventions with greater nuance and depth for specific priority populations (e.g., transgender women) or specific types of structural interventions (e.g., structural interventions targeting patient access). We will share out results at research conferences. Finally, we will incorporate the results into a literature review dashboard (https://hivimpsci.northwestern.edu/dashboard/) our team has created out of previous systematic reviews. This dashboard will provide a high-level visualization of determinants, implementation strategies, and adjunctive interventions by CFIR, ERIC, the TDF and COM-B, as well as stratified by HIV EBI, structural intervention type, recipient/patient population, provider role(s), delivery setting, geographic region of implementation, and publication year. The dashboard is meant to serve researchers in exploring what has already been identified by prior research, to aid them in deciding the direction of future research, as well as to assist them in quickly identifying literature to include in publications and grant submissions. Further, the dashboard is meant to serve as a tool to implementation practitioners, who may seek to understand what determinants have been identified in their particular delivery settings or the population(s) they serve and to identify potential implementation strategies or adjunctive interventions to implement in their own delivery settings.

### Timeline

We have thus far completed the initial search and the NLP screening process. We are currently conducting title and abstract review. Full text review and extraction will begin January 2025 with both analyses beginning June 2025 and running through June 2026. Dissemination will occur in Fall 2025 through Fall 2026.

## Discussion

Our systematic review aims to address the critical gap in understanding and targeting the barriers and facilitators to implementing HIV structural interventions in the U.S and will provide insights that are essential for guiding efforts to scale up HIV structural interventions. Understanding these implementation determinants, implementation strategies, and adjunctive interventions is crucial for reaching marginalized populations who continue to face disparities in access to and engagement with HIV EBIs.

Social and economic determinants such as housing stability, transportation, and healthcare access are critical factors influencing HIV outcomes, particularly for racially, sexually, and gender minoritized groups, and those with co-occurring conditions including substance misuse, serious mental illness, and other infectious diseases and chronic conditions which make treatment engagement more challenging (4,6,19). The implementation strategies and adjunctive interventions to be identified in this review can inform future policy and program development. For example, strategies that leverage digital health technologies, patient transportation and housing, and adaptation of existing EBIs could address systemic barriers such as geographic, financial, and social/linguistic constraints.

## Conclusion

By providing a comprehensive analysis of the implementation determinants, implementation strategies, and adjunctive interventions for HIV structural interventions, our review will identify existing gaps in the literature and provide researchers with an understanding of the needs of the field. These findings hold the potential to significantly advance the goals of the Ending the HIV Epidemic initiative, particularly by addressing the complex needs of populations that have historically been left behind in the fight against HIV.

## Data Availability

All data produced in the present study are available upon reasonable request to the authors.

## List of abbreviations

CFIR: The Consolidated Framework for Implementation Research
EBI: Evidence based intervention
ERIC: Expert Recommendations for Implementation Change
GBMSM: Gay, bisexual, and other men who have sex with men
IS: Implementation science
NLP: Natural language processing
PrEP: Pre-exposure prophylaxis
PRISMA: Preferred reporting items for systematic reviews and meta-analyses
SDOH: Social and structural determinants of health
TDF: Theoretical Domains Framework

## Declarations

### Ethics approval and consent to participate

Not applicable.

### Consent for publication

Not applicable

### Availability of data and materials

The datasets used and/or analysed during the current study are available from the corresponding author on reasonable request.

## Acknowledgments

The authors would like to thank James Merle who assisted us in designing the NLP screening strategy. The authors would also like to thank Corinne Miller who provided consultation on the search strategy.

