## Supplemental File 1 for "Surveying the Literature on Implementation Determinants and Strategies for HIV Structural Interventions: A Systematic Review Protocol"

Supplemental File 1
Search Strategy

| Database | Search | Results |
|---|---|---|
| PubMed (via OVID MEDLINE) | (Acquired Immunodeficiency Syndrome[mesh] OR HIV[mesh] OR AIDS[mesh] OR Human immunodeficiency virus[Title/Abstract] OR AIDS-related[Title/Abstract] OR HIV infection*[mesh] OR pre-exposure prophyla*[Title/Abstract] OR anti-retroviral agent*[Title/Abstract] OR anti-retroviral therap*[Title/Abstract] OR antiretroviral therap*[Title/Abstract] OR antiretroviral treatment[Title/Abstract] OR ART[mesh] OR antiretroviral agent*[Title/Abstract] OR PrEP[mesh] OR post-exposure prophyla*[Title/Abstract] OR PEP[Title/Abstract] OR nPEP[Title/Abstract]) AND (Quality Improvement[mesh] OR Implement*[Title/Abstract] OR Program evaluation[publication type] OR Evaluat*[Title/Abstract] OR "Delivery of Health Care"[Title/Abstract] OR Delivery Model[Title/Abstract] OR Behavioral Science[mesh] OR Social Science[mesh]) AND (structural intervention*[mesh] OR decriminaliz*[Title/Abstract] OR policy[Title/Abstract] OR policies[Title/Abstract] OR policy intervention[Title/Abstract] OR structural change mechanism[Title/Abstract] OR hous*[Title/Abstract] OR employ*[Title/Abstract] OR educat*[Title/Abstract] OR train*[Title/Abstract] OR empower*[Title/Abstract] OR syringe exchange[Title/Abstract] OR harm reduc*[Title/Abstract] OR gender affirm*[Title/Abstract] OR criminaliz*[Title/Abstract] OR financ*[Title/Abstract] OR invest*[Title/Abstract] OR centraliz*[Title/Abstract] OR community mobiliz*[Title/Abstract] OR social market*[Title/Abstract] OR mass media[Title/Abstract] OR government*[Title/Abstract] OR legislati*[Title/Abstract] OR integrat*[Title/Abstract] OR fund*[Title/Abstract] OR survival[Title/Abstract] OR stigma reduction[Title/Abstract] OR reduce stigma[Title/Abstract] OR reducing stigma[Title/Abstract] OR social intervention[Title/Abstract] OR health equity[Title/Abstract] OR equity[Title/Abstract] OR equality[Title/Abstract] OR homophobi*[Title/Abstract] OR transphobi*[Title/Abstract] OR "social determinants of health"[Title/Abstract] OR social determinant*[Title/Abstract] OR structural determinant*[Title/Abstract] OR "structural determinants of health"[Title/Abstract] OR contingency management[Title/Abstract] OR antiracis*[Title/Abstract] OR capacity building[Title/Abstract] OR health | 2,440 |

| | | |
|---|---|---|
| | policy[Title/Abstract] OR needle-exchange[Title/Abstract] OR needle exchange[Title/Abstract] OR socioeconomic[Title/Abstract] OR racis*[Title/Abstract] OR social stigma[Title/Abstract] OR law[Title/Abstract] OR legal[Title/Abstract] OR ordinance*[Title/Abstract] OR statue*[Title/Abstract] OR regulat*[Title/Abstract] OR rule*[Title/Abstract]) AND "2000/01/01"[Date - Publication] : "3000"[Date - Publication] AND English[Language]) AND (English[Language] NOT (“Developing Countries”[Mesh] OR "Africa"[Mesh] OR "Europe"[Mesh] OR "South America"[Mesh] OR "Canada"[Mesh] OR "Greenland"[Mesh] OR "Mexico"[Mesh] OR "Asia"[Mesh] OR "Oceania"[Mesh] OR "Central America"[Mesh] OR “Latin America”[MeSh] OR "Low and Middle Income Countries"[Title/Abstract] OR LMIC[Title/Abstract]) AND United States[mesh] | |
| Embase (Elsevier) | (Acquired Immunodeficiency Syndrome/exp OR HIV/exp OR AIDS/exp OR Human immunodeficiency virus:Ab,ti OR AIDS-related:Ab,ti OR HIV infection/exp OR pre-exposure prophyla*Ab,ti OR anti-retroviral agent*Ab,ti OR anti-retroviral therap*:Ab,ti OR antiretroviral therap*:Ab,ti OR antiretroviral treatment:Ab,ti OR ART/exp OR antiretroviral agent*:Ab,ti OR PrEP/exp OR post-exposure prophyla*:Ab,ti OR PEP:Ab,ti OR nPEP:Ab,ti) AND (Quality Improvement/exp OR Implement*Ab,ti OR Program evaluation:/exp OR Evaluat*:Ab,ti OR Delivery of Health Care:Ab,ti OR Delivery Model:Ab,ti OR Behavioral Science/exp OR Social Science/exp) AND (structural intervention*/exp OR decriminaliz*:Ab,ti OR policy:Ab,ti OR policies:Ab,ti OR policy intervention:Ab,ti OR structural change mechanism:Ab,ti OR hous*:Ab,ti OR employ*:Ab,ti OR educat*:Ab,ti OR train*:Ab,ti OR empower*:Ab,ti OR syringe exchange:Ab,ti OR harm reduc*:Ab,ti OR gender affirm*:Ab,ti OR criminaliz*:Ab,ti OR financ*:Ab,ti OR invest*:Ab,ti OR centraliz*:Ab,ti OR community mobiliz*:Ab,ti OR social market*:Ab,ti OR mass media:Ab,ti OR government*:Ab,ti OR legislati*:Ab,ti OR integrat*:Ab,ti OR fund*:Ab,ti OR survival:Ab,ti OR stigma reduction:Ab,ti OR reduce stigma:Ab,ti OR reducing stigma:Ab,ti OR social intervention:Ab,ti OR health equity:Ab,ti OR equity:Ab,ti OR equality:Ab,ti OR homophobi*:Ab,ti OR transphobi*:Ab,ti OR social determinants of health:Ab,ti OR social determinant*:Ab,ti OR structural determinant*:Ab,ti OR structural determinants of health:Ab,ti OR contingency management:Ab,ti OR antiracis*:Ab,ti OR capacity building:Ab,ti OR health policy:Ab,ti OR needle-exchange:Ab,ti OR needle | 51 |

| | | |
|---|---|---|
| | exchange:Ab,ti OR socioeconomic:Ab,ti OR racis*:Ab,ti OR social stigma:Ab,ti OR law:Ab,ti OR legal:Ab,ti OR ordinance*:Ab,ti OR statue*:Ab,ti OR regulat*:Ab,ti OR rule*:Ab,ti) NOT (Developing Countries OR Africa OR Europe OR South America OR Canada OR Greenland OR Mexico OR Asia OR Oceania OR Central America OR Latin America OR Low and Middle Income Countries:Ab,ti OR LMIC:Ab,ti) AND United States | |
| PsycINFO (EbscoHost) | (ALL="Acquired Immunodeficiency Syndrome" OR ALL=HIV OR ALL=AIDS OR (TI="Human immunodeficiency virus" OR AB="Human immunodeficiency virus") OR (TI=AIDS-related OR AB=AIDS-related) OR ALL="HIV infection*" OR (TI="pre-exposure prophyla*" OR AB="pre-exposure prophyla*") OR (TI="anti-retroviral agent*" OR AB="anti-retroviral agent*") OR (TI="anti-retroviral therap*" OR AB="anti-retroviral therap*") OR (TI="antiretroviral therap*" OR AB="antiretroviral therap*") OR (TI="antiretroviral treatment" OR AB="antiretroviral treatment") OR ALL=ART OR (TI="antiretroviral agent*" OR AB="antiretroviral agent*") OR ALL=PrEP OR (TI="post-exposure prophyla*" OR AB="post-exposure prophyla*") OR (TI=PEP OR AB=PEP) OR (TI=ncep OR AB=ncep)) AND (ALL="Quality Improvement" OR (TI=Implement* OR AB=Implement*) OR ALL="Program evaluation" OR (TI="Delivery of Health Care" OR AB="Delivery of Health Care") OR ALL="Behavioral Science" OR ALL="Social Science") AND (ALL="structural intervention*" OR (TI=decriminaliz* OR AB=decriminaliz*) OR (TI=policy OR AB=policy) OR (TI=policies OR AB=policies) OR (TI="policy intervention" OR AB="policy intervention") OR (TI="structural change mechanism" OR AB="structural change mechanism") OR (TI=hous* OR AB=hous*) OR (TI=employ* OR AB=employ*) OR (TI=educat* OR AB=educat*) OR (TI=train* OR AB=train*) OR (TI=empower* OR AB=empower*) OR (TI="syringe exchange" OR AB="syringe exchange") OR (TI="harm reduc*" OR AB="harm reduc*") OR (TI="gender affirm*" OR AB="gender affirm*") OR (TI=criminaliz* OR AB=criminaliz*) OR (TI=financ* OR AB=financ*) OR (TI=invest* OR AB=invest*) OR (TI=centraliz* OR AB=centraliz*) OR (TI="community mobiliz*" OR AB="community mobiliz*") OR (TI="social market*" OR AB="social market*") OR (TI="mass media" OR AB="mass media") OR (TI=government* OR AB=government*) OR (TI=legislati* OR AB=legislati*) OR (TI=integrat* OR AB=integrat*) OR (TI=fund* OR | 48 |

| | | |
|---|---|---|
| | AB=fund*) OR (TI=survival OR AB=survival) OR (TI="stigma reduction" OR AB="stigma reduction") OR (TI="reduce stigma" OR AB="reduce stigma") OR (TI="reducing stigma" OR AB="reducing stigma") OR (TI="social intervention" OR AB="social intervention") OR (TI="health equity" OR AB="health equity") OR (TI=equity OR AB=equity) OR (TI=equality OR AB=equality) OR (TI=homophobi* OR AB=homophobi*) OR (TI=transphobi* OR AB=transphobi*) OR (TI="social determinants of health" OR AB="social determinants of health") OR (TI="social determinant*" OR AB="social determinant*") OR (TI="structural determinant*" OR AB="structural determinant*") OR (TI="structural determinants of health" OR AB="structural determinants of health") OR (TI="contingency management" OR AB="contingency management") OR (TI=antiracis* OR AB=antiracis*) OR (TI="capacity building" OR AB="capacity building") OR (TI="health policy" OR AB="health policy") OR (TI=needle-exchange OR AB=needle-exchange) OR (TI="needle exchange" OR AB="needle exchange") OR (TI=socioeconomic OR AB=socioeconomic) OR (TI=racis* OR AB=racis*) OR (TI="social stigma" OR AB="social stigma") OR (TI=law OR AB=law) OR (TI=legal OR AB=legal) OR (TI=ordinance* OR AB=ordinance*) OR (TI=statue* OR AB=statue*) OR (TI=regulat* OR AB=regulat*) OR (TI=rule* OR AB=rule*)) NOT (ALL="Developing Countries" OR ALL=Africa OR ALL=Europe OR ALL="South America" OR ALL=Canada OR ALL=Greenland OR ALL=Mexico OR ALL=Asia OR ALL=Oceania OR ALL="Central America" OR ALL="Latin America" OR (TI="Low and Middle Income Countries" OR AB="Low and Middle Income Countries") OR (TI=LMIC OR AB=LMIC)) AND ALL="United States" and Article (Document Types) and Article (Document Types) and English (Languages) and USA (Countries/Regions) \| 5,078 results | |
| Web of Science | (ALL="Acquired Immunodeficiency Syndrome" OR ALL=HIV OR ALL=AIDS OR (TI="Human immunodeficiency virus" OR AB="Human immunodeficiency virus") OR (TI=AIDS-related OR AB=AIDS-related) OR ALL="HIV infection*" OR (TI="pre-exposure prophyla*" OR AB="pre-exposure prophyla*") OR (TI="anti-retroviral agent*" OR AB="anti-retroviral agent*") OR (TI="anti-retroviral therap*" OR AB="anti-retroviral therap*") OR (TI="antiretroviral therap*" OR AB="antiretroviral therap*") OR (TI="antiretroviral treatment" OR AB="antiretroviral | 5,162 |

| | |
|---|---|
| | treatment") OR ALL=ART OR (TI="antiretroviral agent*" OR AB="antiretroviral agent*") OR ALL=PrEP OR (TI="post-exposure prophyla*" OR AB="post-exposure prophyla*") OR (TI=PEP OR AB=PEP) OR (TI=ncep OR AB=ncep)) AND (ALL="Quality Improvement" OR (TI=Implement* OR AB=Implement*) OR ALL="Program evaluation" OR (TI="Delivery of Health Care" OR AB="Delivery of Health Care") OR ALL="Behavioral Science" OR ALL="Social Science") AND (ALL="structural intervention*" OR (TI=decriminaliz* OR AB=decriminaliz*) OR (TI=policy OR AB=policy) OR (TI=policies OR AB=policies) OR (TI="policy intervention" OR AB="policy intervention") OR (TI="structural change mechanism" OR AB="structural change mechanism") OR (TI=hous* OR AB=hous*) OR (TI=employ* OR AB=employ*) OR (TI=educat* OR AB=educat*) OR (TI=train* OR AB=train*) OR (TI=empower* OR AB=empower*) OR (TI="syringe exchange" OR AB="syringe exchange") OR (TI="harm reduc*" OR AB="harm reduc*") OR (TI="gender affirm*" OR AB="gender affirm*") OR (TI=criminaliz* OR AB=criminaliz*) OR (TI=financ* OR AB=financ*) OR (TI=invest* OR AB=invest*) OR (TI=centraliz* OR AB=centraliz*) OR (TI="community mobiliz*" OR AB="community mobiliz*") OR (TI="social market*" OR AB="social market*") OR (TI="mass media" OR AB="mass media") OR (TI=government* OR AB=government*) OR (TI=legislati* OR AB=legislati*) OR (TI=integrat* OR AB=integrat*) OR (TI=fund* OR AB=fund*) OR (TI=survival OR AB=survival) OR (TI="stigma reduction" OR AB="stigma reduction") OR (TI="reduce stigma" OR AB="reduce stigma") OR (TI="reducing stigma" OR AB="reducing stigma") OR (TI="social intervention" OR AB="social intervention") OR (TI="health equity" OR AB="health equity") OR (TI=equity OR AB=equity) OR (TI=equality OR AB=equality) OR (TI=homophobi* OR AB=homophobi*) OR (TI=transphobi* OR AB=transphobi*) OR (TI="social determinants of health" OR AB="social determinants of health") OR (TI="social determinant*" OR AB="social determinant*") OR (TI="structural determinant*" OR AB="structural determinant*") OR (TI="structural determinants of health" OR AB="structural determinants of health") OR (TI="contingency management" OR AB="contingency management") OR (TI=antiracis* OR AB=antiracis*) OR (TI="capacity building" OR AB="capacity building") OR (TI="health policy" OR AB="health policy") OR (TI=needle-exchange OR |

| | | |
|---|---|---|
| | AB=needle-exchange) OR (TI="needle exchange" OR AB="needle exchange") OR (TI=socioeconomic OR AB=socioeconomic) OR (TI=racis* OR AB=racis*) OR (TI="social stigma" OR AB="social stigma") OR (TI=law OR AB=law) OR (TI=legal OR AB=legal) OR (TI=ordinance* OR AB=ordinance*) OR (TI=statue* OR AB=statue*) OR (TI=regulat* OR AB=regulat*) OR (TI=rule* OR AB=rule*)) NOT (ALL="Developing Countries" OR ALL=Africa OR ALL=Europe OR ALL="South America" OR ALL=Canada OR ALL=Greenland OR ALL=Mexico OR ALL=Asia OR ALL=Oceania OR ALL="Central America" OR ALL="Latin America" OR (TI="Low and Middle Income Countries" OR AB="Low and Middle Income Countries") OR (TI=LMIC OR AB=LMIC)) AND ALL="United States" and Article (Document Types) and Article (Document Types) and English (Languages) and USA (Countries/Regions) \| 5,078 results | |
| Sociological Abstracts | ((Acquired Immunodeficiency Syndrome) OR (HIV) OR (AIDS) OR title(Human immunodeficiency virus) OR abstract(Human immunodeficiency virus) OR title(AIDS-related) OR abstract(AIDS-related) OR (hiv infections) OR title(pre-exposure prophylaxis) OR abstract(pre-exposure prophylaxis) OR title(anti-retroviral agent) OR abstract(anti-retroviral agent) OR title(anti-retroviral therapy) OR abstract(anti-retroviral therapy) OR title(antiretroviral therapy) OR abstract(antiretroviral therapy) OR title(antiretroviral treatment) OR abstract(antiretroviral treatment) OR (ART) OR abstract(antiretroviral agents) OR abstract(antiretroviral agents) OR (PrEP) OR title(post-exposure prophylaxis) OR abstract(post-exposure prophylaxis) OR title(PEP) OR abstract(PEP) OR title(nPEP) OR abstract(nPEP)) AND ((Quality Improvement) OR title(Implement) OR title(implementation) OR title(implementating) OR abstract(Implement) OR abstract(implementation) OR abstract(implementing) OR (Program evaluation) OR title(Delivery of Health Care) OR abstract(Delivery of Health Care) OR (Behavioral Science) OR (Social Science)) AND ((structural interventions) OR title(decriminalize) OR title(decriminalization) OR abstract(decriminalize) OR abstract(decriminalization) OR title(policy) OR abstract(policy) OR title(policies) OR abstract(policies) OR title(policy intervention) OR abstract(policy intervention) OR title(structural change mechanism) OR abstract(structural change mechanism) OR title(housing) OR abstract(housing) OR title(employ) OR title(employment) OR abstract(employ) OR | 191 |

| | |
|---|---|
| | abstract(employment) OR title(educate) OR title(educating) OR title(education) OR abstract(educate) OR abstract(educating) OR abstract(education) OR title(train) OR title(training) OR abstract(train) OR abstract(training) OR title(empower) OR title(empowering) OR title(empowerment) OR abstract(empower) OR abstract(empowerment) OR abstract(empowering) OR title(syringe exchange) OR abstract(syringe exchange) OR title(harm reduction) OR abstract(harm reduction) OR title(gender affirming) OR title(gender affirmation) OR abstract(gender affirming) OR abstract(gender affirmation) OR title(criminalize) OR title(criminalizing) OR abstract(criminalize) OR abstract(criminalizing) OR title(financing) OR abstract(financing) OR title(invest) OR title(investing) OR title(investment) OR abstract(invest) OR abstract(investing) OR abstract(investment) OR title(centralize) OR title(centralizing) OR abstract(centralize) OR abstract(centralizing) OR title(community mobilization) OR abstract(community mobilization) OR title(social market) OR title(social marketing) OR abstract(social market) OR abstract(social marketing) OR title(mass media) OR abstract(mass media) OR title(government) OR abstract(government) OR title(legislate) OR title(legislation) OR title(legislating) OR abstract(legislate) OR abstract(legislation) OR abstract(legislating) OR title(integrate) OR title(integration) OR title(integrating) OR abstract(integrate) OR abstract(integrating) OR abstract(integration) OR title(fund) OR title(funding) OR abstract(fund) OR abstract (funding) OR title(survival) OR abstract(survival) OR title(stigma reduction) OR abstract(stigma reduction) OR title(reduce stigma) OR abstract(reduce stigma) OR title(reducing stigma) OR abstract(reducing stigma) OR title(social intervention) OR abstract(social intervention) OR title(health equity) OR abstract(health equity) OR title(equity) OR abstract(equity) OR title(equality) OR abstract(equality) OR title(homophobia) OR title(homophobic) OR abstract(homophobia) OR abstract(homophobic) OR title(transphobia) OR title(transphobic) OR abstract(transphobia) OR abstract(transphobic) OR title(social determinants of health) OR abstract(social determinants of health) OR title(social determinants) OR abstract(social determinants) OR title(structural determinant) OR abstract(structural determinan) OR title(structural determinants of health) OR abstract(structural determinants of health) OR title(contingency management) OR abstract(contingency management) OR title(antiracism) OR title(antiracist) OR |

| | | |
|---|---|---|
| | abstract(antiracism) OR abstract(antiracist) OR title(capacity building) OR abstract(capacity building) OR title(health policy) OR abstract(health policy) OR title(needle-exchange) OR abstract(needle-exchange) OR title(needle exchange) OR abstract(needle exchange) OR title(socioeconomic) OR abstract(socioeconomic) OR title(racism) OR title(racist) OR abstract(racism) OR abstract(racist) OR title(social stigma) OR abstract(social stigma) OR title(law) OR abstract(law) OR title(legal) OR abstract(legal) OR title(ordinance) OR abstract(ordinance) OR title(statute) OR abstract(statute) OR title(regulate) OR title(regulation) OR abstract(regulate) OR abstract(regulation) OR title(rule) OR abstract(rule)) NOT ((Developing Countries) OR (Africa) OR (Europe) OR (South America) OR (Canada) OR (Greenland) OR (Mexico) OR (Asia) OR (Oceania) OR (Central America) OR (Thailand) OR (Kenya) OR (Brazil) OR (Latin America) OR title(Low and Middle Income Countries) OR abstract(Low and Middle Income Countries) OR title(LMIC) OR abstract(LMIC)) | |
| Google | ("structural intervention"\|"structural interventions")(HIV\|AIDS\|"immunodeficiency virus") site:.gov | 18 (9 automatically removed bc they were funding announcements, 1 removed bc textbook, 3 being a policy document with no analysis, 2 duplicate, 2 non-domestic) |
| ProQuest Congressional CRS Reports and Miscellaneous Publications | ("structural intervention" OR "structural interventions") AND (HIV OR "Human Immunodeficiency Virus" OR "acquired immune deficiency syndrome--aids") | 16 |
